# Disulfiram associated with lower risk of Covid-19: a retrospective cohort study

**DOI:** 10.1101/2021.03.10.21253331

**Authors:** Nathanael Fillmore, Steven Bell, Ciyue Shen, Vinh Nguyen, Jennifer La, Maureen Dubreuil, Judith Strymish, Mary Brophy, Gautam Mehta, Hao Wu, Judy Lieberman, Nhan Do, Chris Sander

**Author notes:** joint first authors (Nathanael Fillmore, Steven Bell, Ciyue Shen). joint senior authors (Nhan Do, Chris Sander).

## Abstract

In the global COVID-19 pandemic, there is a substantial need for effective, low-cost therapeutics. We investigated the potential effects of disulfiram on the incidence and outcomes of COVID-19 in an observational study in a large database of US Veterans Administration clinical records, the VA Corporate Data Warehouse (CDW). The study is motivated by the unique properties of disulfiram, which has been used as an anti-alcoholism drug since 1948, is non-toxic, easy to manufacture and inexpensive. Disulfiram reduces hyperinflammation in mammalian cells by inhibition of the gasdermin D pore. In a mouse model of sepsis, disulfiram reduced inflammatory cytokines and mortality. Disulfiram also is a low micromolar inhibitor of the M^pro^ and PL^pro^ viral proteases of SARS-CoV-2.

To investigate the potential effects of disulfiram on the incidence and severity of COVID-19, we carried out an epidemiological study in the CDW. The VA dataset used has 944,127 patients tested for SARS-Cov-2, 167,327 with a positive test, and 2,233 on disulfiram, of which 188 had a positive SARS-Cov-2 test. A multivariable Cox regression adjusted for age, gender, race/ethnicity, region, a diagnosis of alcohol use disorders, and Charlson comorbidity score revealed a reduced incidence of COVID-19 with disulfiram use with a hazard ratio of **0.66** and 95% confidence interval of 0.57 to 0.76 (P < 0.001). There were no deaths among the 188 SARS-Cov-2 positive patients treated with disulfiram. The expected number of deaths would have been 5-6 according to the 3% death rate among the untreated (P-value 0.03).

Our finding of a lower hazard ratio and less severe outcomes for COVID-19 in patients treated with disulfiram compared to those not treated is a statistical association and does not prove any causative effect of disulfiram. However, the results of this study suggest that there is a pharmacological contribution to the reduced incidence and severity of COVID-19 with the use of disulfiram. Given the known anti-inflammatory and viral anti-protease effects of disulfiram, it is reasonable and urgent to initiate accelerated clinical trials to assess whether disulfiram reduces SARS-CoV-2 infection, disease severity and death.

**STRUCTURED OUTLINE:** *Importance:* Identifying already approved medications with well characterized antiviral or anti-inflammatory properties supported by real world evidence as candidates for clinical trials for repurposing is an important strategy to manage the pandemic given the ongoing challenges with producing and administering vaccines, the emergence of more infectious viral mutants and the paucity of approved therapies.

*Objective:* To investigate the potential effects of disulfiram on the incidence and severity of COVID-19.

*Design:* Retrospective cohort study from February 20, 2020 to February 1, 2021.

*Setting:* Veterans Health Administration. Veterans who had visited a VA primary care provider in the 18 months before their first SARS-CoV-2 test.

*Participants:* 2,233 Veterans with at least one SARS-CoV-2 laboratory (positive or negative) test result on or after February 20, 2020 and at least one pharmacy record for disulfiram on or after February 20, 2019 and 941,894 Veterans without a pharmacy record for disulfiram.

*Exposure:* Treatment with disulfiram

*Main Outcome:* Positive test result for SARS-CoV-2

*Results:* A multivariable Cox regression analysis adjusted for age, gender, race/ethnicity, region, diagnosis of an alcohol use disorder, and Charlson comorbidity score resulted in a reduced hazard of COVID-19 infection with disulfiram use, with a hazard ratio of 0.66 and 95% confidence interval of 0.57 to 0.76 (P < 0.001).

*Conclusions and Relevance:* The results of this study suggest that disulfiram use contributes to a reduced incidence of COVID-19. Given the known anti-inflammatory and anti-protease effects of disulfiram, its low cost, low side effects, and general availability, it is reasonable and urgent to initiate accelerated clinical trials to assess the effect of disulfiram on infection and the development of advanced disease.

## INTRODUCTION

### The need for effective treatment of Covid-19

There have been more than 100 million cases of coronavirus disease 2019 (COVID-19) worldwide since the beginning of the pandemic, with over 2.3 million deaths. Although several vaccines against severe acute respiratory syndrome coronavirus 2 (SARS-Cov-2) have been approved and are being administered internationally, there will still be a significant number of infections in people who are not vaccinated or for whom vaccination fails to fully protect. In addition, there will likely be significant delays in vaccinating many people worldwide due to difficulties in producing and administering vaccinations in some settings (resource poor countries). In light of this, identifying effective, inexpensive, and widely accessible treatment strategies is a high priority.

### Repurposing approved drugs

Given the length of time and associated costs involved in developing new drugs, repurposing of medications that are already approved for another indication is an attractive strategy that has been utilized for other diseases (1). With a growing number of candidates being identified, conducting a clinical trial for each remains a challenge due to the substantial time and financial resources required. While not equivalent to a randomized controlled study (so any conclusions drawn are correlations, not proven causative effects) real world data can be used to investigate the incidence or severity of disease in those prescribed a medication of interest compared to those not taking the drug to help prioritize prospective therapeutic targets by providing observational evidence of a drug’s potential effectiveness against COVID-19 (1).

### Focus on disulfiram

This study focuses on providing such data for disulfiram (sold under the trade name Antabuse), an aldehyde dehydrogenase inhibitor used to treat alcohol use disorder (2), with over 60 years of clinical use and an acceptable safety profile. This is motivated both by promising observations about the effects of disulfiram in several biochemical and cell-based screens as well as in mouse experiments (3–12).

### Molecular mechanisms of disulfiram

The potential antiviral effects of disulfiram are primarily a reduction of hyperinflammation in host cells and inhibition of virus-encoded proteases. There are at least four lines of evidence to support this. **(1) Anti-inflammatory effects**: based on the results of a drug screen of 3,752 compounds, Hu *et al*. (3) discovered a pronounced anti-inflammatory effect of disulfiram and reduction of mortality in a mouse model of sepsis, suggesting a possible therapeutic effect in severe COVID-19 disease. Disulfiram inhibits inflammatory cytokine IL-1β release and pyroptosis (inflammatory cell death) by blocking the assembly of the 27-subunit 108-beta-stranded homopolymeric gasdermin D pore by covalent modification of Cys191. **(2) Inhibition of the main protease:** Jin *et al*. (4) conducted high throughput screening of potential inhibitors of the SARS-CoV-2 main protease (M^pro^, also known as 3CL^pro^ or 3-chymotrypsin-like protease), an enzyme needed for viral replication and identified disulfiram among the candidates. They measured an IC50 of 9.35 μM for disulfiram, which may be physiologically relevant given the dosage of disulfiram prescribed for alcohol use disorders (up to 500mg daily). **(3) Inhibition of the papain-like protease:** earlier Lin et al. (5) demonstrated the inhibition of the papain-like proteases (PL^pro^) of MERS-CoV and SARS-CoV-1 by disulfiram through multiple enzymatic assays. The inhibition of PL^pro^ of the SARS-CoV-2 virus has been confirmed by several groups, at IC50 values in biochemical assays with purified protein ranging from 2-7 μM ((6–8) and Robert Davey, private communication). The inhibition of PL^pro^ is reported to be the result of disruption of a Cys_4_-Zinc ion binding site and/or via covalent modification of the active site Cys residue (8). However, these results may not translate directly to an *in vivo* inhibitory effect, as the formation of S-S covalent bonds between disulfiram and its target Cys residues strongly depends on reducing conditions (6,10). **(4) Inhibition of other viral proteins:** it has also been reported that disulfiram can inhibit the ATPase activity of nsp13 and the exoribonuclease activity of nsp14 of SARS-CoV-2 via its Zn^2+^-ejector function (11). These mechanistic studies support the hypothesis that disulfiram may be a polypharmacological therapeutic compound in coronavirus disease.

### Aim of this study

In this study, we look for additional evidence to be considered when prioritizing existing medications for clinical trials in both the early and late stages of COVID-19. We investigate whether there are significant differences in rates of incidence or in the outcome of COVID-19 disease among persons taking disulfiram compared to the general population.

## METHODS

### Data and Patients

We conducted a retrospective cohort study comparing Veterans who received treatment with disulfiram to those who were not treated, based on data from the Department of Veterans Affairs (VA) COVID-19 Shared Data Resource and the VA Corporate Data Warehouse (CDW), which consolidates electronic health record data from VA facilities nationwide. We identified a cohort of Veterans with at least one SARS-CoV-2 laboratory test result between February 20, 2020 and February 1, 2021. In addition, in order to ensure patients were receiving routine care at the VA, we also required that each Veteran had visited a VA primary care provider in the 18 months before their first SARS-CoV-2 test. Within this cohort, we identified 2,233 Veterans (median age 51 years [IQR 39, 61], 8.4% female) with at least one pharmacy record for disulfiram between February 20, 2019 and February 1, 2021, and 941,894 Veterans (median age 64 years [IQR 51, 72], 11.6% female) not prescribed disulfiram between those dates. Disulfiram pharmacy records were identified in the CDW based on standardized generic drug names.

### Study Variables and Outcomes

The primary outcome of interest was the occurrence of a positive test result for SARS-CoV-2 (either PCR or antigen), as recorded in the VA COVID-19 Shared Data Resource. Patients were followed from February 20, 2020 through the date of a positive SARS-CoV-2 test, the date of death, or the end of the study (February 1, 2021), whichever came first. Date of death was defined based on mortality records available in the CDW, integrating data from Medicare, Social Security Administration, VA facilities, National Cemetery Administration, and death certificates (13). In addition to the primary outcome, we also considered severe clinical outcomes subsequent to SARS-CoV-2 infection, including death, intensive care unit (ICU) admission, and mechanical ventilation occurring within 2 weeks of a positive SARS-CoV-2 test, as defined in our previous work (14). For a composite severe endpoint, we combined death, ICU admission, and mechanical ventilation.

Study variables included age at start of follow-up (i.e., February 20, 2020), gender, race/ethnicity, region of the VA facility where each patient was tested for SARS-CoV-2, diagnosis of alcohol use disorder (AUD), and the Charlson comorbidity index. Age, gender, race, and ethnicity were defined based on structured data in the CDW. The region of the VA facility where each patient was tested for SARS-CoV-2 was determined using a mapping previously reported (14). Diagnosis of AUD in the year prior to the start of follow-up was determined based on an algorithm adapted for VA from the Centers for Medicare and Medicaid Chronic Conditions Warehouse algorithms (15). The Charlson comorbidity index was calculated using the R *comorbidity* package based on ICD-10 codes recorded in the year prior to the start of follow-up (16).

### Statistical Analysis

We summarized patients’ demographics at baseline, i.e., measured at or before the start of follow-up, for the full analytic sample and by disulfiram treatment status. We also tested for baseline differences by treatment status using t-tests for continuous variables and chi-squared tests for categorical variables.

We conducted two sets of analyses to examine the associations between disulfiram treatment and SARS-CoV-2 test positivity. First, we calculated the incidence rate of SARS-CoV-2 infection during follow-up time before and after disulfiram treatment, and assessed differences using the incidence rate ratio and a Wald test. Patients were considered to have been treated with disulfiram during all follow-up time on or after the date of their first pharmacy record for a dispensed disulfiram prescription, as in an intention-to-treat analysis. If the patient received disulfiram between February 20, 2019 and February 20, 2020, all follow-up time was coded as post-treatment. If the positive test occurred during follow-up, the follow-up time before the first pharmacy record was coded as pre-treatment and time after that record was coded as post-treatment. For the untreated cohort, if the patient did not have any disulfiram pharmacy records, all follow-up time was coded as pre-treatment.

Second, we fit univariable and multivariable Cox proportional hazards models, adjusting for covariates listed in **Table 1**. In multivariable analysis, age was included as a continuous variable, measured in years, while other variables were categorical. While the primary approach assumed that those who were treated with disulfiram continued to be exposed throughout the remaining follow up time, we also performed a sensitivity analysis considering that patients remained on disulfiram for only one month after each record of dispensation. Another sensitivity analysis was restricted to those with diagnosis of AUD. Analyses were conducted using R v4.0.2 (http://www.r-project.org) and p-values < 0.05 were considered statistically significant.

**Table 1.** Patient characteristics in the full analytic sample and stratified by patients who did and did not receive disulfiram in the study period. Notation for number of patients: N (%); IQR = Interquartile range.

| Patient Characteristics | Full Analytic Sample | Never Treated with Disulfiram | Treated with Disulfiram |
| --- | --- | --- | --- |
| **All patients** | 944127 | 941894 (99.8) | 2233 (0.2) |
| **Age (median [IQR])** | 64 [51, 72] | 64 [51, 72] | 51 [39, 61] |
| **Gender (%)** |  |  |  |
| Male | 834982 (88.4) | 832936 (88.4) | 2046 (91.6) |
| Female | 109144 (11.6) | 108957 (11.6) | 187 (8.4) |
| **Race/Ethnicity (%)** |  |  |  |
| Non-Hispanic White | 573612 (60.8) | 571928 (60.7) | 1684 (75.4) |
| Non-Hispanic Black | 204379 (21.6) | 204130 (21.7) | 249 (11.2) |
| Hispanic | 80154 (8.5) | 80025 (8.5) | 129 (5.8) |
| Other or Unknown | 85982 (9.1) | 85811 (9.1) | 171 (7.7) |
| **Region (%)** |  |  |  |
| Continental | 154710 (16.4) | 154449 (16.4) | 261 (11.7) |
| Midwest | 197872 (21.0) | 197216 (20.9) | 656 (29.4) |
| North Atlantic | 213784 (22.6) | 213214 (22.6) | 570 (25.5) |
| Pacific | 178755 (18.9) | 178310 (18.9) | 445 (19.9) |
| Southeast | 199004 (21.1) | 198703 (21.1) | 301 (13.5) |
| **Charlson Score (%)** |  |  |  |
| 0 | 361778 (38.3) | 360708 (38.3) | 1070 (47.9) |
| 1-2 | 373407 (39.6) | 372503 (39.5) | 904 (40.5) |
| 3-4 | 132843 (14.1) | 132657 (14.1) | 186 (8.3) |
| >=5 | 50689 (5.4) | 50644 (5.4) | 45 (2.0) |
| Unknown | 25410 (2.7) | 25382 (2.7) | 28 (1.3) |
| **History of AUD (%)** | 100873 (10.7) | 98910 (10.5) | 1963 (87.9) |
| **Positive Covid-19 test** | 167327 (17.7) | 167139 (17.7) | 188 (8.4) |

### Institutional Review Board

This study was approved by the institutional review board at VA Boston Healthcare System (IRB3328).

## RESULTS

### The VA patient cohort

Among 944,127 patients meeting the inclusion criteria, 2,233 had at least one pharmacy record for disulfiram on or after February 20, 2019. The patient population is racially and regionally heterogeneous, reflecting the VA’s nationwide patient population (**Table 1**). Patients treated with disulfiram differed in their basic demographic and clinical history from those who were not treated, including being younger in age, more likely to be white, and having a lower burden of comorbidity (P < 0.001 for all tests).

### Lower COVID-19 incidence with disulfiram

The incidence rate of SARS-CoV-2 infection in patients treated with disulfiram was 112.3 per 1000 person years of follow-up (95% confidence interval [CI] 96.8 - 129.5), based on 188 patients infected during 1,674 person years of follow-up. In contrast, the incidence rate of infection in untreated patients was 199.4 per 1000 person years of follow-up (95% CI 198.4 - 200.3), based on 167,139 patients infected during 838,279 person years of follow-up. Reflecting this difference, the **incidence rate ratio** of infection in treated vs untreated patients was **0.56** (95% CI 0.49 - 0.65, P < 0.001), i.e., a 44% lower rate of infection in those treated with disulfiram.

### Lower COVID-19 hazard with disulfiram

In the **univariable** model, we observed a hazard ratio (HR) of **0.50** (95% CI 0.44 - 0.58, P < 0.001), similar to the incidence rate ratio, as expected. In the **multivariable** model (Figure 1), after adjusting for age, gender, race/ethnicity, region, Charlson score, and diagnosis of an alcohol use disorder, the association was still highly significant. Patients treated with disulfiram had a 34% lower risk of SARS-CoV-2 infection compared to those not treated with disulfiram, with a hazard ratio of **0.66** (95% CI 0.57 - 0.76, P < 0.001).

**Figure 1.**
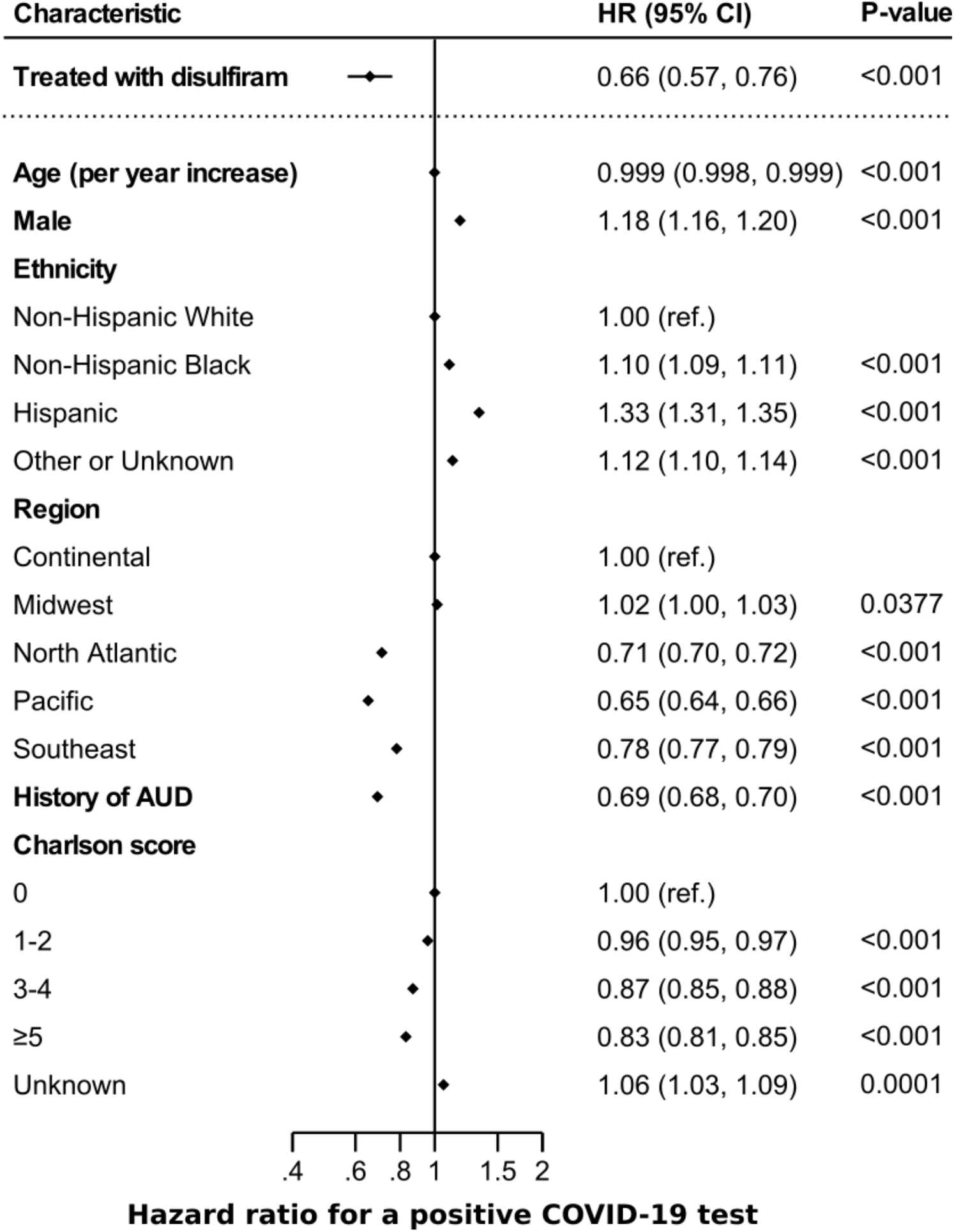
Hazard ratios (HR) and 95% confidence intervals (CI) for a positive SARS-Cov-2 test for patients treated with disulfiram compared to those not treated with disulfiram; results from a multivariable Cox model in an unrestricted analysis of all 944127 patients, adjusting for covariates measured at baseline, i.e., measured at or before the start of follow-up. Disulfiram is coded as a time-dependent covariate. AUD: Alcohol Use Disorder

### Alternative Analyses

For the multivariable model sensitivity analysis, assuming that patients remained on disulfiram for only one month after each record of dispensation, we observed a similar association with a hazard ratio of 0.53 (95% CI 0.44 - 0.64, P <0.001). In the analysis restricted to those with diagnosis of AUD (100,873 out of 944,127 patients), we observed a hazard ratio of **0.68** (95% CI 0.58 - 0.80, P < 0.001) (Supplementary Table S1 and Figure S1). Within confidence limits, this result is consistent with the hazard ratio of 0.66 (previous paragraph) of the unrestricted analysis.

### Assessing factors contributing to disease risk

The multivariable model also revealed significant associations of risk of infection with age, gender, race/ethnicity, region, and comorbidity (via the Charlson score), which are consistent with earlier results (14). The lower risk of infection with a higher Charlson score might be due to an effect of comorbidities on reduction of social contacts. We cannot resolve this potential effect, as data on social contacts is not available. However, the multivariable analysis does provide an estimate of the extent to which the association of reduced incidence with disulfiram use is confounded by other factors.

### COVID-19 outcomes

Beyond the incidence of disease, we also report counts of severe clinical outcomes and a composite measure of severe outcomes (Table 2). Inferences from these data are limited due to the low frequency of events. There appear to be no statistically significant differences in ICU admissions and mechanical ventilation between the two cohorts. We did observe that no deaths were reported among the 188 patients in the disulfiram cohort, while 5-6 deaths would have been expected according to the 3% death rate in the untreated cohort (Table 2). Overall, there was not a statistical significant difference between the proportion of the composite severe endpoints between the two cohorts. Given the low counts of severe clinical outcomes in this study and uncertainty regarding disulfiram concentration level after discontinuation on admission, epidemiological observations in a larger dataset or in an inpatient setting, such as in clinical trials, are needed to establish the effects of disulfiram on the severity of COVID-19.

**Table 2.** Clinical outcomes in patients infected with SARS-Cov-2 by disulfiram treatment status. P values for the significance of distinction between treated and untreated.

| **Outcome** | No. with Outcome<br>Treated | Total No.<br>Treated | No. with Outcome<br>Untreated | Total No.<br>Untreated | Incidence<br>Proportion<br>Treated | Incidence<br>Proportion<br>Untreated | P |
| --- | --- | --- | --- | --- | --- | --- | --- |
| **ICU** | 11 | 188 | 7,403 | 167,139 | 0.059 | 0.044 | 0.442 |
| **Mechanical**<br>**Ventilation** | 1 | 188 | 959 | 167,139 | 0.005 | 0.006 | 1 |
| **Death** | 0 | 188 | 5,009 | 167,139 | **0** | **0.030** | 0.028 |
| **All Severe**<br>**Outcomes** | 12 | 188 | 13,371 | 167,139 | 0.063 | 0.080 | 0.447 |

## DISCUSSION

### Comparison with earlier work

While we recognize the risk of residual confounding variables with the observational nature of the study design, our study is the largest using real world data to demonstrate a lower risk of SARS-CoV-2 infection in those taking disulfiram. Recently, in a preliminary report, Tamburin *et al*. (17) showed individuals taking disulfiram had a significantly lower risk of COVID-19 compatible symptoms, such as fever and dyspnea, compared to a control group. However their study was underpowered to reliably discern a difference in risk for a positive COVID-19 test, rather than just for compatible symptoms, between the groups. Our study significantly extends these initial observations in two ways: (1) the VA patient population with relevant data is considerably larger, which enabled us to infer a statistically significant association between disulfiram use and risk of test-certified COVID-19 infection, and (2) we were able to document the clinical outcomes of those infected with COVID-19 taking disulfiram, although our study was underpowered to establish a statistically significant correlation between disulfiram use and less severe outcomes. Access to larger clinical datasets and meta-analysis across diverse health-care systems would be highly beneficial in future investigations on this topic.

### Limitations of the study

The principal limitation of our study, as is true even for studies with controlled cohorts, is that association does not prove causation and that even very likely causation does not guarantee the success of the corresponding therapeutic intervention. There are several additional limitations. Since Veterans can receive care outside of the VA, diagnoses and outcomes may be incompletely recorded; and, we do not know if patients actually took the prescribed disulfiram. We also cannot rule out the possibility of confounding by indication: that those prescribed disulfiram had other factors, such as behavioral differences that contributed to the protection against COVID-19 beyond the effect of the medication.

### The potential of clinical trials of disulfiram in COVID-19

Our results reinforce the results from candidate drug screens and in-depth molecular and physiological evidence in supporting the notion that disulfiram may be efficacious in combating COVID-19. Additional information is expected from two small ongoing Phase II clinical trials of disulfiram involving early mild-to-moderate symptomatic (NCT04485130, 60 participants) or hospitalized (NCT04594343, 200 participants) COVID-19 patients. Reduction in disease progression in the *first trial*, if observed, might be due to inhibition of viral replication. This hypothesis is based on clear evidence from biochemical experiments that disulfiram inhibits several enzymes involved in viral replication (Mpro, PLpro, nsp13 and nsp14) via covalent modification of cysteine residues in the active site of the Mpro and PLpro proteases and via weakening of Cys-cordinated Zn2+ ion binding sites in PLpro and in the RNA-modifying enzymes nsp13 and nsp14 (4–12). Better outcomes in the *second trial*, if observed, might be due to the anti-inflammatory effects of disulfiram, as demonstrated in mouse models of sepsis, via inhibition of the formation of the host gasdermin pore and reduction of pyroptosis (3).

### Potential impact on current and future COVID disease

Given the urgent medical need, additional clinical trials in uninfected populations as well as in early and late COVID-19 disease are highly desirable with the aim of testing the impact that disulfiram may have across the entire COVID-19 continuum, from infection to disease progression and including the current and constantly evolving SARS-CoV-2 variants. As a repurposing candidate, disulfiram has particular advantages given its low cost, both in production and distribution, and favorable effect profile (2,18).

We propose that clinical trials should be pursued pro-actively, given the current limited access to vaccination and the lack of generally available, low-cost proven therapeutics against the disease caused by the original SARS-CoV-2 virus and its variants. Our view is that such trials require governmental or philanthropic funding and that time is of the essence. If successful in clinical trials, disulfiram may be a good candidate as a general anti-COVID-19 therapeutic for world-wide distribution, including to low-income populations.

## Data Availability

US Veterans Administration clinical records are protected information in the VA Corporate Data Warehouse (CDW). Only summary statistics are reported in the manuscript.

## ACKNOWLEDGEMENTS

We thank Soeren Brunak at the University of Copenhagen, Sjoerd van Hagen at The Hyve in the Netherlands, Robert Davey at Boston University, Debora Marks at Harvard Medical School and Guy Montelione at Rensselaer Polytechnic for discussions. The work was supported by the British Heart Foundation RG/4/32218 (SB), the VA Boston Healthcare System (VA team), Dana-Farber Cancer Institute and Harvard Medical School (C.Shen, C.Sander).

## SUPPLEMENTARY MATERIALS

**Supplementary Table S1.** Patient characteristics in the **restricted** analysis. The restriction is to the 100873 patients who have a diagnosis of AUD, stratified by patients who did and did not receive disulfiram in the study period. Notation for number of patients: N (%); IQR = Interquartile range.

| Patient Characteristics | Full Analytic Sample | Never Treated with Disulfiram | Treated with Disulfiram |
| --- | --- | --- | --- |
| **All patients** | 100873 | 98910 | 1963 |
| **Age (median [IQR])** | 60 [47, 68] | 60 [48, 68] | 51 [39, 60] |
| **Gender (%)** |  |  |  |
| Male | 93631 (92.8) | 91813 (92.8) | 1818 (92.6) |
| Female | 7242 (7.2) | 7097 (7.2) | 145 (7.4) |
| **Race/Ethnicity (%)** |  |  |  |
| Non-Hispanic White | 56366 (55.9) | 54888 (55.5) | 1478 (75.3) |
| Non-Hispanic Black | 28022 (27.8) | 27804 (28.1) | 218 (11.1) |
| Hispanic | 8897 (8.8) | 8782 (8.9) | 115 (5.9) |
| Other or Unknown | 7588 (7.5) | 7436 (7.5) | 152 (7.7) |
| **Region (%)** |  |  |  |
| Continental | 16142 (16.0) | 15921 (16.1) | 221 (11.3) |
| Midwest | 21896 (21.7) | 21322 (21.6) | 574 (29.2) |
| North Atlantic | 23630 (23.4) | 23129 (23.4) | 501 (25.5) |
| Pacific | 18034 (17.9) | 17637 (17.8) | 397 (20.2) |
| Southeast | 21171 (21.0) | 20901 (21.1) | 270 (13.8) |
| **Charlson Score (%)** |  |  |  |
| 0 | 38724 (38.4) | 37799 (38.2) | 925 (47.1) |
| 1-2 | 40641 (40.3) | 39816 (40.3) | 825 (42.0) |
| 3-4 | 15002 (14.9) | 14831 (15.0) | 171 (8.7) |
| >=5 | 6413 (6.4) | 6372 (6.4) | 41 (2.1) |
| Unknown | 93 (0.1) | 92 (0.1) | 1 (0.1) |
| **Positive Covid-19 test** | 13189 | 13025 | 164 |

**Supplementary Figure S1.**
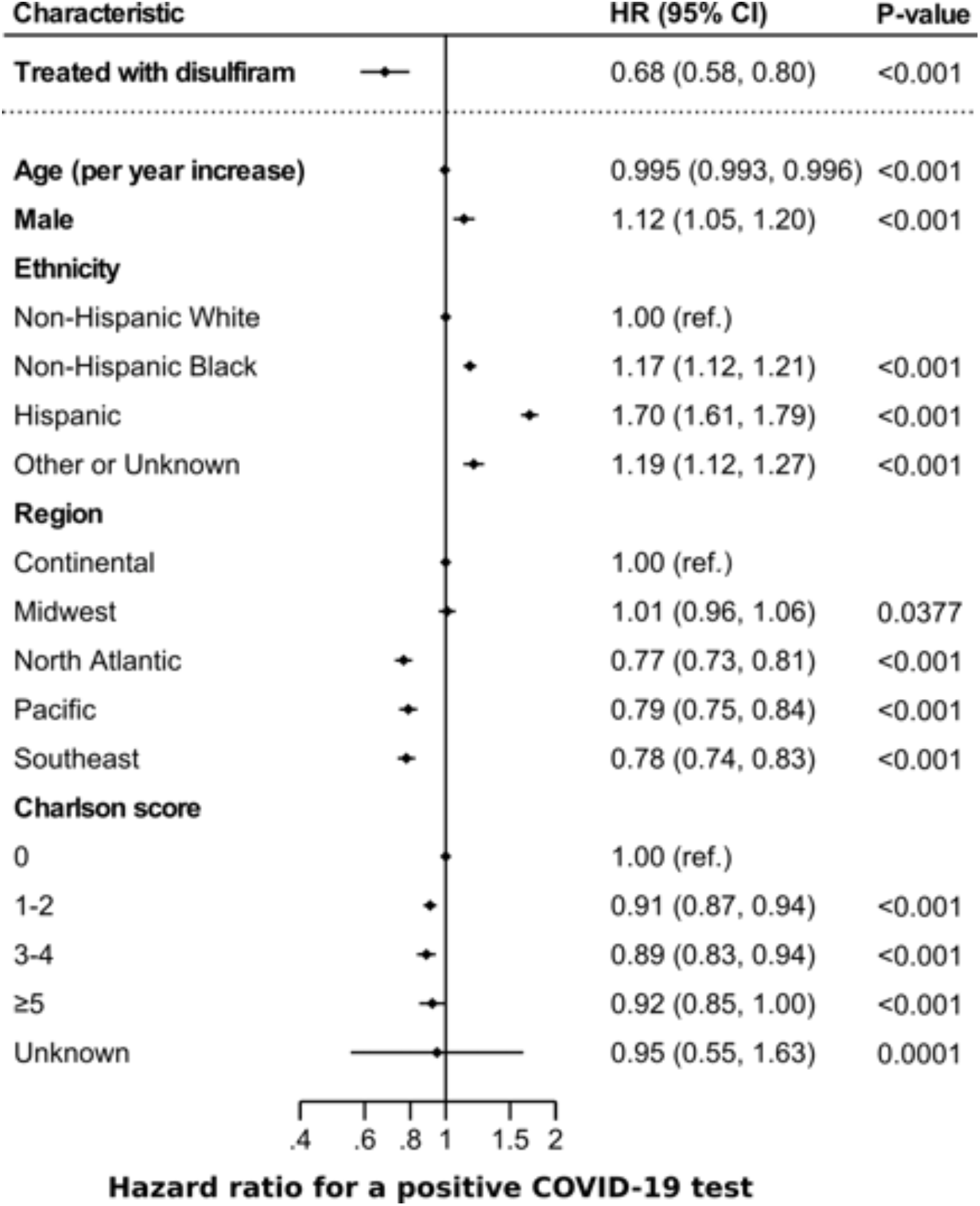
Hazard ratios (HR) for a positive COVID-19 test for patients treated with disulfiram compared to those not treated with disulfiram, in a **restricted** analysis of only patients with AUD. Results from a multivariable Cox model adjusting for covariates measured at baseline, i.e., measured at or before the start of follow-up. Disulfiram is coded as a time-dependent covariate. CI = 95% confidence intervals.

